# Dental Composite Performance Prediction Using Artificial Intelligence

**DOI:** 10.1101/2024.10.08.24314998

**Authors:** Karla Paniagua Rivera, Kyumin Whang, Krishna Joshi, Hyeonwi Son, Yu Shin Kim, Mario Flores

## Abstract

**Objective:** There is a need to increase the performance and longevity of dental composites and accelerate the translation of novel composites to the market. This study explores artificial intelligence (AI), specifically machine learning (ML), to predict the performance outcomes (POs) of dental composites from their composite attributes (CAs).

**Methods:** An extensive dataset from over 200 publications was built and refined to 233 samples with 17 CAs and 7 POs. Nine ML models were evaluated for PO prediction performance using classified data, and Five ML models were evaluated for PO regression analysis.

**Results:** The KNN model excelled in predicting flexural modulus (FlexMod), Decision Tree model in flexural strength (FlexStr) and volumetric shrinkage (ShrinkV), and Logistic Regression and SVM models in shrinkage stress (ShrinkStr). Receiver operating characteristic area under the curve (ROC AUC) analysis confirmed these results but found that Random Forest was more effective for FlexStr and ShrinkV, suggesting the possibility of Decision Tree overfitting the data. Regression analysis revealed that the Voting Regressor was superior for FlexMod and ShrinkV predictions, while Decision Tree Regression was optimal for FlexStr and ShrinkStr. Feature importance analysis indicated TEGDMA is a key contributor to FlexMod and ShrinkV, BisGMA and UDMA to FlexStr, and depth of cure, degree of monomer-to-polymer conversion, and filler loading to ShrinkStr.

**Significance:** There is a need to conduct a full analysis using multiple ML models because different models predict different POs better, and for a large, comprehensive dataset to train robust AI models to facilitate the prediction and optimization of composite properties and support the development of new dental materials.

## 1. INTRODUCTION

At the 2019 Minamata Convention on Mercury, the World Dental Federation and International Association for Dental Research urged for more research on alternatives so dental amalgam restorations can be phased out [1]. This is because amalgam restorations still have significantly higher survivability and longevity than composites [2]. Composite success rates have been reported at 85% over 55 months compared to 92% for amalgam, and longevity is estimated at <8 years [3–5]. Main causes of composite failure include polymerization shrinkage stress and resin degradation leading to marginal gap and secondary caries formation, and restoration fracture [4, 6]. Thus, a longer-lasting restorative is an urgent oral health need.

There have been numerous publications on the development of novel dental composites [7–14]. However, most commercial composites still use the classic bisphenol A glycidyl dimethacrylate (BisGMA), ethoxylated bisphenol A dimethacrylate (BisEMA), urethane dimethacrylate (UDMA) and triethylene glycol dimethacrylate (TEGDMA) and glass filler combination due to the time, effort and expense needed to receive FDA approval for new monomers and filler. Also, modifying material composition to improve composite properties requires repetitive and inefficient *in vitro* formulation and testing. And since the relationship between composition and material properties is non-linear and multifactorial, it is difficult to design an optimal formulation for a specific material property.

Data driven technologies and artificial intelligence (AI), especially Machine Learning (ML) and Deep Learning (DL), are intriguing methods for computational analyses in many fields, including computational material science [15]. DL models are better at tackling non-linear classification tasks and understanding the contributions of each component and what the model is learning. Thus, there is an explosive growth in the literature on ML and DL methods.

In dentistry, AI has been used to diagnose periodontally compromised teeth [16] and periodontal disease [17], predict dental implant success rate [17], make decisions to extract teeth for orthodontic treatment [18], etc., and was often found be as effective as trained specialists [19].

In dental materials, at the time of this writing, there was only one publication on using AI, and that was to determine the effect of composite attributes (CA) on the flexural strength of CAD/CAM resin composite blocks (RCB). UDMA, TEGDMA, and filler content were most important in predicting strength with high prediction performance and low error [20]. However, the utility of this information is limited because 1) only monomer type, filler type and loading were used and variables like monomer concentration were not (probably because commercial composite formulations are trade secrets); 2) a single source (manufacturer values or one article) was used for each product, which may be the reason for the high prediction performance and low error, i.e. the model may be overfitting the data; 3) the sample size of 12 is extremely small for AI models; 4) CAD/CAM composites are more homogeneous than direct composites; and, 5) CAD/CAM RCBs are not nearly as widely used as direct restorative composites. Furthermore, this model may not be as useful for direct composites, which would be subject to many more variables, such as the method of cure, curing lamp used, curing time, etc. Thus, a more extensive literature search for direct dental composites showed a large range of strength values (performance outcome, PO) for the same product. Finally, other POs need to be assessed since strength is not the only important PO for composites and other POs are needed to better determine how effective the AI is in identifying the most important CAs and optimizing composite design.

Thus, the overall objective is to investigate the efficacy of AI models in determining the effects of CAs and predicting composite POs to aid in the development of durable composites and reduce the time, work and expense needed to translate experimental composites to the market. Specifically, for this work, our goal was to build an extensive and unique dataset of CAs and POs and determine the efficacy of different ML models in predicting composite POs.

## 2. MATERIALS AND METHODS

### 2.1. Dataset Construction

Google Scholar (scholar.google.com) was used to identify scientific articles on dental composites. Company brochures were also used to further find composite attributes (CAs) and composite performance outcomes (POs). The CAs gathered were monomer type, monomer concentration, monomer molecular weight, initiators used, initiator concentrations, degree of monomer-to-polymer conversion, filler type, filler loading, filler shape, filler size, viscosity, density, and index of refraction. Composite POs gathered were depth of cure (which could also be a CA), radiopacity, flexural modulus, flexural strength, compressive strength, fracture toughness, fracture work, polymerization volumetric shrinkage, polymerization shrinkage stress, wear depth and cycles, water sorption, water solubility, survival rate, survival length, in vitro cytotoxicity (ED50), total fluoride ion release, ion release duration, and remineralization. This information was recorded in separate columns in a Microsoft Excel sheet, as available. This data was then preprocessed before being analyzed using nine AI models.

### 2.2. Data Preprocessing

All string values were changed to either integer or floating values, depending on its type. Before performing the AI analysis for each PO, samples that did not have values for that PO were removed and from the remaining set of samples, CA’s that only had 0’s were removed and not included in the analysis. Missing values in the dataset’s CAs were imputed with KNN imputation for regression analysis [21], and multivariate imputation for classification analysis [22, 23]. By imputing the missing values, we enhanced the dataset to be as complete and representative as possible, maximizing the amount of usable data, preserving the statistical properties of the dataset and ensuring robust and reliable results. All further analyses were performed on the imputed data. Finally, since the data is continuous, each PO datum was classified and divided into two different classes (high/low) with respect to its mean value, where low means that the PO value is smaller or equal to the mean value, and high, means the PO value is larger than the mean value.

### 2.3. AI Analysis

After data preprocessing, the curated dataset was small, so (1) feature engineering was used to extract meaningful features from the limited data; (2) simple ML models that can work with little data were used to prevent overfitting, which would give an artificially and incorrectly high accuracy; and, (3) evaluation tools were implemented to assess the model’s performance.

#### 2.3.1. Classification

For classification, nine different ML classification algorithms were trained using 80% of the curated and imputed data, including Support Vector Machine (SVM), Decision Tree, KNeighbors Classifier (KNN), Light Gradient Boosting Machine (LGBM), Random Forest, Logistic Regression, Gaussian Naïve Bayes, Extreme Learning Machine (ELM), and Extreme Grading Boosting (XGBoost). The remaining 20% of the data was used to test the AI models on how well the models predicted each composite PO. To evaluate the classification models’ performances, evaluation tools, such as accuracy (the proportion of correctly classified instances), precision (the proportion of true positive instances out of all instances predicted as positive), recall (the proportion of true positive instances out of all actual positive instances), and F1score (the harmonic mean of precision and recall, i.e. how well those two correlate) were used.

The Receiver Operating Characteristic Area Under the Curve (ROC AUC) and the AUC scores were also computed for each model. The ROC AUC curve plots the true positive rate (sensitivity) against the false positive rate (1-specificity) to show the model’s performance, and the higher the AUC scores the better the model’s performance [24].

The dataset was further analyzed using the best performing model with a feature importance tool to find and rank the importance of features (CAs) in increasing or decreasing each PO: Decision Tree, Random Forest, XGBoost or LGBM [25]. Feature importance tool measures the contribution of each CA to the model’s predictions by evaluating how much each CA reduces impurity in the decision-making process. In tree architecture models, impurity refers to how mixed the data is at a particular node in the tree in terms of the target class (Figure S1). High impurity occurs when a node has a mix of different classes, which makes it harder to classify the data. Conversely, low impurity occurs when a node contains mostly one class, indicating more homogeneous and better-classified data. Note that results do not specify whether the CA increases or decreases the PO, just how important it is in changing the PO.

Permutation importance was also determined. This method randomly shuffles the values of each feature (CA), observes its impact on the model’s accuracy and provides an estimate of its importance in the model’s accuracy in predicting low or high PO. So, while the overall feature importance calculated above shows the contribution of each CA to increasing or decreasing the PO, permutation importance shows how important the CA is for the model to correctly predict the classification of the PO.

#### 2.3.2. Regression Analysis

Since the original data is continuous, five regression models were trained on 80% of the curated and imputed data subset and tested on the remaining 20% to predict those continuous values. The five models used were: Support Vector Regression (SVR) [26, 27], Decision Tree Regression [28], Histogram Gradient Boosting Regression (HistGradientBoost) [29], Random Forest Regression [30], and a voting regressor (an ensemble learning method that combines the predictions from multiple regression models by averaging the individual predictions from each model to produce a final prediction with improved accuracy and robustness). The voting regressor is composed of Linear Regression, Random Forest Regression, KNN regression, SVR, HistGradientBoost, and Decision Tree regression.

To evaluate the regression models’ performances, the explained variance score, R^2^, was calculated. It measures the variance in the PO as well as the influence of the CAs on these fluctuations in the model and quantifies how well the model accounts for the variability. Additionally, how well a model predicts the POs was assessed by calculating five different errors: Mean Absolute Error (MAE), Mean Squared Error (MSE), Root Mean Squared Error (RMSE), Median Absolute Error (MedAE), and Max Error (ME). These errors ensures that different aspects of the model’s predictive accuracy are evaluated and facilitates model selection, refinement and interpretation of results: MAE provides an average of absolute errors [31], MSE emphasizes larger errors [31], RMSE gives a measure in the original units of the target variable [31], MedAE is robust against outliers [32], and ME highlights the worst-case prediction error [33].

## 3. RESULTS

### 3.1. Dataset Construction

200+ publications and company brochures were used to build the database of CAs and POs. The initial dataset consisted of 321 samples (composites) with 28 CAs and 17 POs. Despite the large number of publications and brochures used, many cells were empty, much of the information for commercial products came from brochures, which are not as trustworthy, and multiple sources of data for the same PO for the same composite gave deviant values. This dataset also included experimental monomers or fillers that were not used in other publications or composites. Thus, when the AI analysis was done, their contribution to the POs were disproportionately underweighted. So, the dataset was curated to include only commercial light cure composites to aid in the evaluation of the AI models.

The final curated dataset contained 233 composites. Table 1 lists the 17 CAs and Table 2 lists the 7 POs in this dataset. Table 1 also shows that there are CAs with missing values, so values were imputed into CAs, as described above (Section 2.2), to ensure that the model can accurately learn patterns and relationships without being biased by missing data.

**Table 1.** Composite Attributes (Cas), Sample Sizes and Proportions to All Samples.

| CA | Sample Size | Proportion (%) |
| --- | --- | --- |
| 1. BisGMA Concentration | 39 | 17 |
| 2. UDMA Concentration | 11 | 5 |
| 3. TEGDMA Concentration | 44 | 19 |
| 4. BisEMA Concentration | 0 | 0 |
| 5. DDDMA Concentration | 0 | 0 |
| 6. DX-511 Concentration | 0 | 0 |
| 7. EGDMA Concentration | 0 | 0 |
| 8. EBPDMA Concentration | 0 | 0 |
| 9. BMAEPP Concentration | 1 | 0.4 |
| 10. PEGDMA Concentration | 0 | 0 |
| 11. Silorane Concentration | 0 | 0 |
| 12. TCDUA Concentration | 0 | 0 |
| 13. MDP Concentration | 0 | 0 |
| 14. Filler Loading (Fill) | 176 | 75 |
| 15. Degree of Monomer to Polymer Conversion (DoC) | 55 | 24 |
| 16. Viscosity (Visc) | 12 | 5 |
| 17. Depth of Cure (DepthCure) | 34 | 15 |

**Table 2.** Composite Performance Outcomes (POs), Sample Sizes, Proportions to All Samples, and Whether They Were Analyzed.

| PO | Sample Size | Proportion (%) | Analyzed? |
| --- | --- | --- | --- |
| 1. Flexura Modulus (FlexMod) | 103 | 44.2 | Yes |
| 2. Flexural Strength (FlexStr) | 128 | 54.9 | Yes |
| 3. Compressive Strength (CompStr) | 42 | 18.0 | No |
| 4. Fracture Toughness (FracTough) | 40 | 17.2 | Yes |
| 5. Fracture Work (FracWk) | 9 | 3.9 | No |
| 6. Polymerization Volumetric Shrinkage (ShrinkV) | 106 | 45.5 | Yes |
| 7. Polymerization Shrinkage Stress (ShrinkStr) | 45 | 19.3 | Yes |

When each PO was analyzed, samples with no values for that specific PO were further removed, resulting in the samples sizes shown in Table 2. FlexStr had the largest sample size with 54.9% of all samples and FrackWk had smallest with 3.9%. There was evidence of overfitting when analyzing POs with small sample sizes, so CompStr, FracTough and FracWk were excluded from further analysis. The overfitting shows the importance of having a large sample size for training robust and accurate ML models.

### 3.2. Classification

#### 3.2.1. Flexural Modulus

Table 3 shows the performances of the different AI models for FlexMod. The KNN model performed the best in predicting FlexMod classification (i.e. whether a CA increases or decreases FlexMod), with an accuracy of 0.90, a precision of 0.92, a recall score of 0.90 and a F1 score of 0.90, highlighting the balanced performance of the KNN model in accurately identifying low and high FlexMod instances while minimizing false positives (low FlexMod miss-predicted as high FlexMod) and negatives (high FlexMod miss-predicted as low FlexMod).

**Table 3.** AI Model Performance for Flexural Modulus (FlexMod)

| Metric | SVM | Decision Tree | KNN | LGBM | Rando m Forest | Logistic Regressio n | Gaussia n Naïve Bayes | ELM | XGBoos t |
| --- | --- | --- | --- | --- | --- | --- | --- | --- | --- |
| **Accuracy** | 0.88 | 0.71 | **0.90** | 0.80 | 0.85 | 0.83 | 0.73 | 0.54 | 0.83 |
| **Precision** | 0.90 | 0.74 | **0.92** | 0.84 | 0.89 | 0.85 | 0.77 | 0.57 | 0.88 |
| **Recall** | 0.88 | 0.71 | **0.90** | 0.80 | 0.85 | 0.83 | 0.73 | 0.54 | 0.83 |
| **F1 score** | 0.88 | 0.71 | **0.90** | 0.80 | 0.85 | 0.82 | 0.73 | 0.53 | 0.83 |

Figure 1a shows the ROC AUC curves for each model. The KNN model had the highest AUC score of 0.97, indicating that the model is very good at distinguishing between high and low FlexMod across different probability thresholds and confirming the model’s performance.

**Figure 1.**
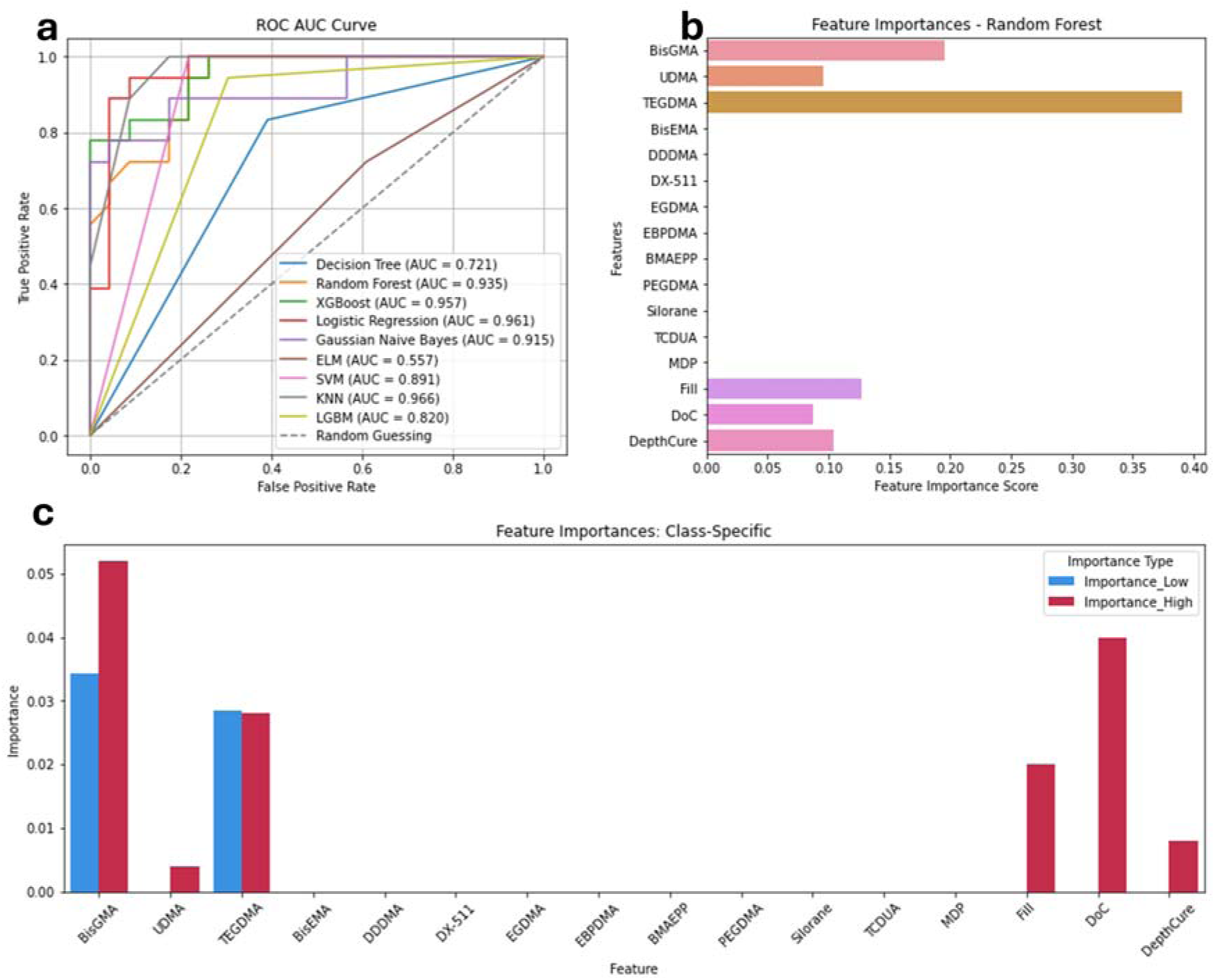
Flexural Modulus. a) ROC AUC Curve of the 9 algorithms, b) Feature importances for Flexural Modulus. c) Feature importances per class.

Figure 1b shows results from the feature importance analysis using the Random Forest model, the highest performing model in Table 3 with a feature importance tool. Random Forest model reduces impurity by finding splits that make nodes purer, resulting in more accurate predictions. The greater the reduction in impurity across all trees in the forest, the higher the importance of the CA. The order of the six CAs that most impact FlexMod from highest to lowest importance is: TEGDMA (Feature Importance Score (FIC) = 0.39), BisGMA (FIC = 0.19), Fill (FIC = 0.13), DepthCure (FIC = 0.10), UDMA (FIC =0.09), and DoC (FIC = 0.08). Other CAs had negligible importance, but that could be due only a few composites containing those CAs. This means, that the model gives more weight to the values in TEDGMA when predicting for FlexMod, followed by BisGMA.

Figure 1c shows the importance of each CA in the ML model predicting whether a composite would have low or high FlexMod. Thus, the permutation importance scores for each CA (Y-axis) indicate how much the model’s predictive accuracy decreases when the values of that CA are randomly shuffled (permuted) for a specific class (low or high FlexMod). If permuting a CA causes a significant drop in predictive accuracy, it means that the feature is crucial for predicting that class. For example, for low FlexMod (using only samples with below average FlexMod), TEGDMA is the most important feature, as permuting this CA presented the largest decrease in the model’s accuracy in predicting low FlexMod, with a drop of 0.048 (4.8%). Similarly, BisGMA is the second most important feature for low FlexMod, with a permutation score (PS) resulting in a 0.04 decrease in accuracy, followed by UDMA (PS = 0.008). For accurately predicting high FlexMod, TEGDMA is also the most important CA (PS = 0.056). Similarly, BisGMA is the second most important feature for predicting high FlexMod (PS = 0.048). Notably, DoC also has a permutation score of 0.048 for high FlexMod. This is followed by Fill (PS = 0.020), DepthCure (PS = 0.008), and UDMA (PS = 0.004). These numbers are low, but since the model uses the scores from all of the features to make predictions, they are still important to note. Thus, when both BisGMA and TEGDMA were permuted simultaneously, the accuracy dropped by 20%. This also means that in the design of composites, the CAs that have the highest PS must be controlled the most to produce composites with low or high FlexMod.

#### 3.2.2. Flexural Strength

For predicting low or high classification of FlexStr, the Decision Tree model performed the best (Table 4), with an accuracy, a precision, a recall score, and an F1 score of 0.73, each. While not high, these numbers are acceptable.

**Table 4.** Performance Scores for Flexural Strength (FlexStr)

| **Metric** | **SVM** | **Decision Tree** | **KNN** | **LGBM** | **Random Forest** | **Logistic Regression** | **Gaussian Naïve Bayes** | **ELM** | **XGBoost** |
| --- | --- | --- | --- | --- | --- | --- | --- | --- | --- |
| **Accuracy** | 0.71 | **0.73** | 0.65 | 0.69 | 0.69 | 0.58 | 0.48 | 0.58 | 0.69 |
| **Precision** | 0.72 | **0.73** | 0.65 | 0.69 | 0.70 | 0.58 | 0.47 | 0.58 | 0.69 |
| **Recall** | 0.71 | **0.73** | 0.65 | 0.69 | 0.69 | 0.58 | 0.48 | 0.58 | 0.69 |
| **F1 score** | 0.71 | **0.73** | 0.65 | 0.69 | 0.69 | 0.57 | 0.43 | 0.58 | 0.69 |

Figure 2a shows the ROC AUC curves for each AI model in predicting FlexStr. Despite the Decision Tree having highest performance scores in Table 4, it only had the third-highest AUC score of 0.73. The Random Forest model had the highest AUC score of 0.802, followed by XGBoost (AUC score = 0.797). This indicates that while the Decision Tree model performed well overall, Random Forest and XGBoost demonstrated superior abilities in distinguishing between low and high FlexStr across various thresholds.

**Figure 2.**
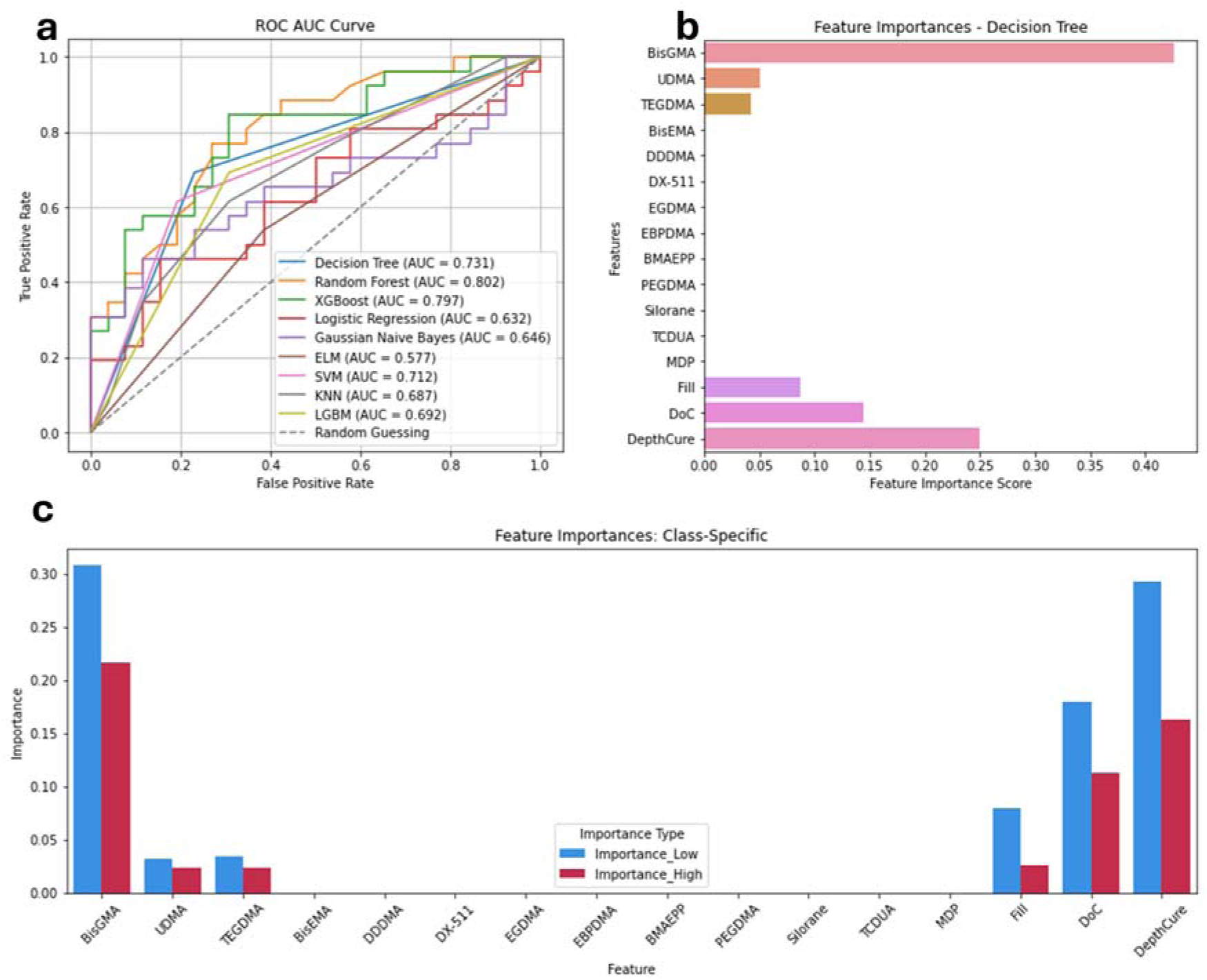
Flexural Strength. a) ROC AUC Curve of the 9 algorithms, b) Feature importances for Flexural Strength. c) Feature importances per class.

Feature importance analysis measured with the Decision Tree model (Figure 2b) showed that the six important CAs for predicting FlexStr in order of highest to lowest importance are: BisGMA (FIS = 0.42), DepthCure (FIS = 0.25), DoC (FIS = 0.14), Fill (FIS = 0.08), UDMA (FIS = 0.05), and TEGDMA (FIS = 0.04). The other CAs had negligible importance.

Figure 2c shows the importance of CAs in predicting low and high FlexStr. For low FlexStr, BisGMA is the most important CA, as permuting this feature in the samples with below-average FlexStr (low classification) presented the largest decrease in the model’s accuracy (PS = 0.31). DepthCure is the second most important feature for classifying low FlexStr (PS = 0.29), followed by DoC (PS = 0.19), Fill (PS = 0.08), TEGDMA (PS = 0.034), and UDMA (PS = 0.031). For high FlexStr we observe a similar pattern (Figure 2c). BisGMA is the most important (PS = 0.22), followed by DepthCure (PS = 0.16), DoC (PS = 0.11), Fill (PS = 0.026), TEGDMA (PS = 0.024), and UDMA (PS = 0.023).

#### 3.2.3. Compressive Strength

Table 5 shows the performance metrics for the AI models in predicting CompStr. There was overfitting with the Decision Tree model, with scores of 1.0, probably due to the limited sample size. Also, in the dataset, of the 17 CAs, only Fill, DoC and DepthCure CAs had values; The rest of the CAs where empty. And from the 42 samples, Fill had 37 values, but DoC had only 11 values and DepthCure 4 values. With only one column having anything close to sufficient data, the models lack enough data to learn from. That leads to generalization, poor performance and overfitting. Imputing missing values may introduce bias or noise, especially in a small dataset, and result in unreliable models. Thus, further analysis was abandoned. More data is needed to ensure robustness and reliability of future analyses of CompStr.

**Table 5.** Performance Scores for Compressive Strength (CompStr)

| Metric | SVM | Decision Tree | KNN | LGBM | Random Forest | Logistic Regression | Gaussian Naïve Bayes | ELM | XGBoost |
| --- | --- | --- | --- | --- | --- | --- | --- | --- | --- |
| **Accuracy** | 0.71 | **1.00** | 0.53 | 0.71 | 0.88 | 0.76 | 0.88 | 0.76 | 0.71 |
| **Precision** | 0.85 | **1.00** | 0.58 | 0.50 | 0.88 | 0.82 | 0.90 | 0.87 | 0.85 |
| **Recall** | 0.71 | **1.00** | 0.54 | 0.71 | 0.88 | 0.76 | 0.88 | 0.77 | 0.71 |
| **F1 score** | 0.72 | **1.00** | 0.55 | 0.58 | 0.88 | 0.70 | 0.87 | 0.77 | 0.72 |

#### 3.2.4. Fracture Toughness

Table 6 shows the performance metrics for the AI models in predicting FracTough. The Random Forest model seems to be overfitting the data, probably due to the limited sample size. Thus, further analysis was abandoned. More data is needed to ensure robustness and reliability of future analyses of FracTough.

**Table 6.**
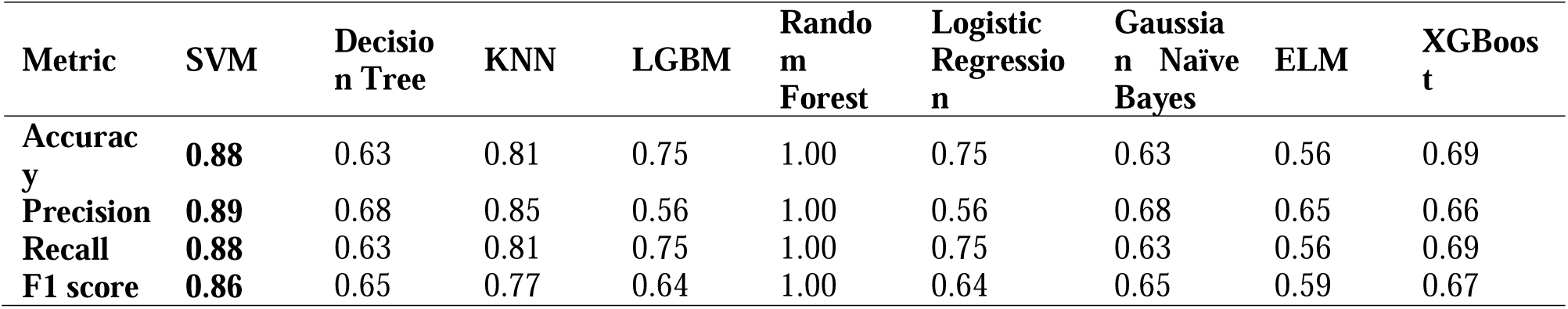
Performance Scores for Fracture Toughness (FracTough)

#### 3.2.5. Volumetric Shrinkage

Table 7 shows that the Decision Tree model had the best performance in predicting ShrinkV classification (accuracy of 0.81, precision of 0.82, recall score of 0.81, and F1 score of 0.82).

**Table 7.** Performance Scores for Polymerization Shrinkage Volume (ShrinkV)

| Metric | SVM | Decision Tree | KNN | LGBM | Random Forest | Logistic Regression | Gaussian Naïve Bayes | ELM | XGBoost |
| --- | --- | --- | --- | --- | --- | --- | --- | --- | --- |
| Accuracy | 0.81 | **0.81** | 0.77 | 0.74 | 0.79 | 0.79 | 0.72 | 0.65 | 0.81 |
| Precision | 0.81 | **0.82** | 0.77 | 0.76 | 0.79 | 0.79 | 0.73 | 0.67 | 0.81 |
| Recall | 0.81 | **0.81** | 0.77 | 0.74 | 0.79 | 0.79 | 0.72 | 0.65 | 0.81 |
| F1 score | 0.81 | **0.82** | 0.77 | 0.75 | 0.79 | 0.78 | 0.72 | 0.66 | 0.81 |

Figure 3a shows the ROC AUC curves for each model in predicting ShrinkV. Despite the Decision Tree having the highest performance in Table 3, it had the fourth-highest AUC score of 0.81. The highest AUC score was achieved by the Random Forest model (AUC score = 0.910), followed by XGBoost (AUC score = 0.889) and Gaussian Naïve Bayes (AUC score = 0.861). This indicates that while the Decision Tree model performed well overall, other models demonstrated superiority in distinguishing between low and high ShrinkV across various thresholds.

**Figure 3.**
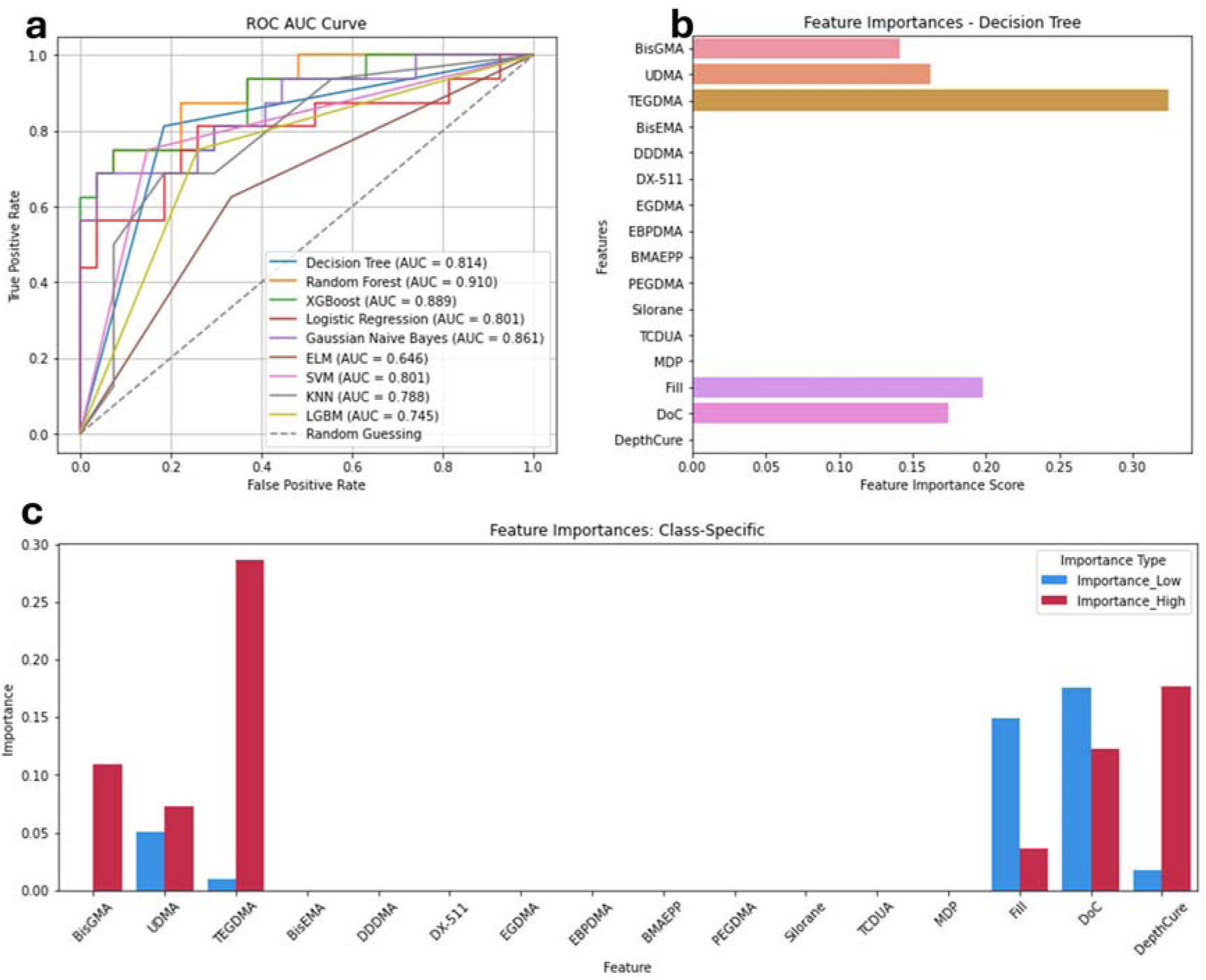
Volumetric Shrinkage. a) ROC AUC Curve of the 9 algorithms, b) Feature importances for Volumetric Shrinkage. c) Feature importances per class.

Feature importance analysis measured with Decision Tree (Figure 3b) showed that the most important CAs for predicting ShrinkV in order of highest to lowest importance were: TEGDMA (FIS = 0.32), Fill (FIS = 0.19), DoC (FIS = 0.17), UDMA (FIS = 0.16), and BisGMA (FIS = 0.14).

Figure 3c shows the importance of CAs in predicting low or high ShrinkV. For low ShrinkV, DoC is the most important CA (PS = 0.17), followed by Fill (PS = 0.14), UDMA (PS = 0.05), DepthCure (PS = 0.17), and TEGDMA (PS = 0.01). For high ShrinkV TEGDMA was the most important CA (PS = 0.28), followed by DepthCure (PS = 0.18), DoC (PS = 0.12), BisGMA (PS = 0.11), UDMA (PS = 0.07), and Fill (PS = 0.04).

#### 3.2.6. Shrinkage Stress

Table 8 shows that Logistic Regression, SVM and XGBoost models had the best performance in predicting ShrinkStr, all with an accuracy of 0.89, a precision of 0.91, a recall score of 0.89, and a F1 score of 0.89.

**Table 8.** Performance scores for Shrinkage Stress classification.

| **Metric** | **SVM** | **Decision Tree** | **KNN** | **LGBM** | **Random Forest** | **Logistic Regression** | **Gaussian Naïve Bayes** | **ELM** | **XGBoost** |
| --- | --- | --- | --- | --- | --- | --- | --- | --- | --- |
| **Accuracy** | **0.89** | 0.56 | 0.78 | 0.50 | 0.61 | **0.89** | 0.83 | 0.61 | **0.89** |
| **Precision** | **0.91** | 0.56 | 0.79 | 0.25 | 0.63 | **0.91** | 0.84 | 0.63 | **0.91** |
| **Recall** | **0.89** | 0.56 | 0.78 | 0.50 | 0.61 | **0.89** | 0.83 | 0.61 | **0.89** |
| **F1 score** | **0.89** | 0.55 | 0.78 | 0.33 | 0.60 | **0.89** | 0.83 | 0.60 | **0.89** |

Figure 4a shows the ROC AUC curves for each model in predicting ShrinkStr. Logistic Regression had the highest AUC score of 0.963), followed by SVM (AUC score = 0.899). However, XGBoost only had an AUC score of 0.444, despite its high-performance scores.

**Figure 4.**
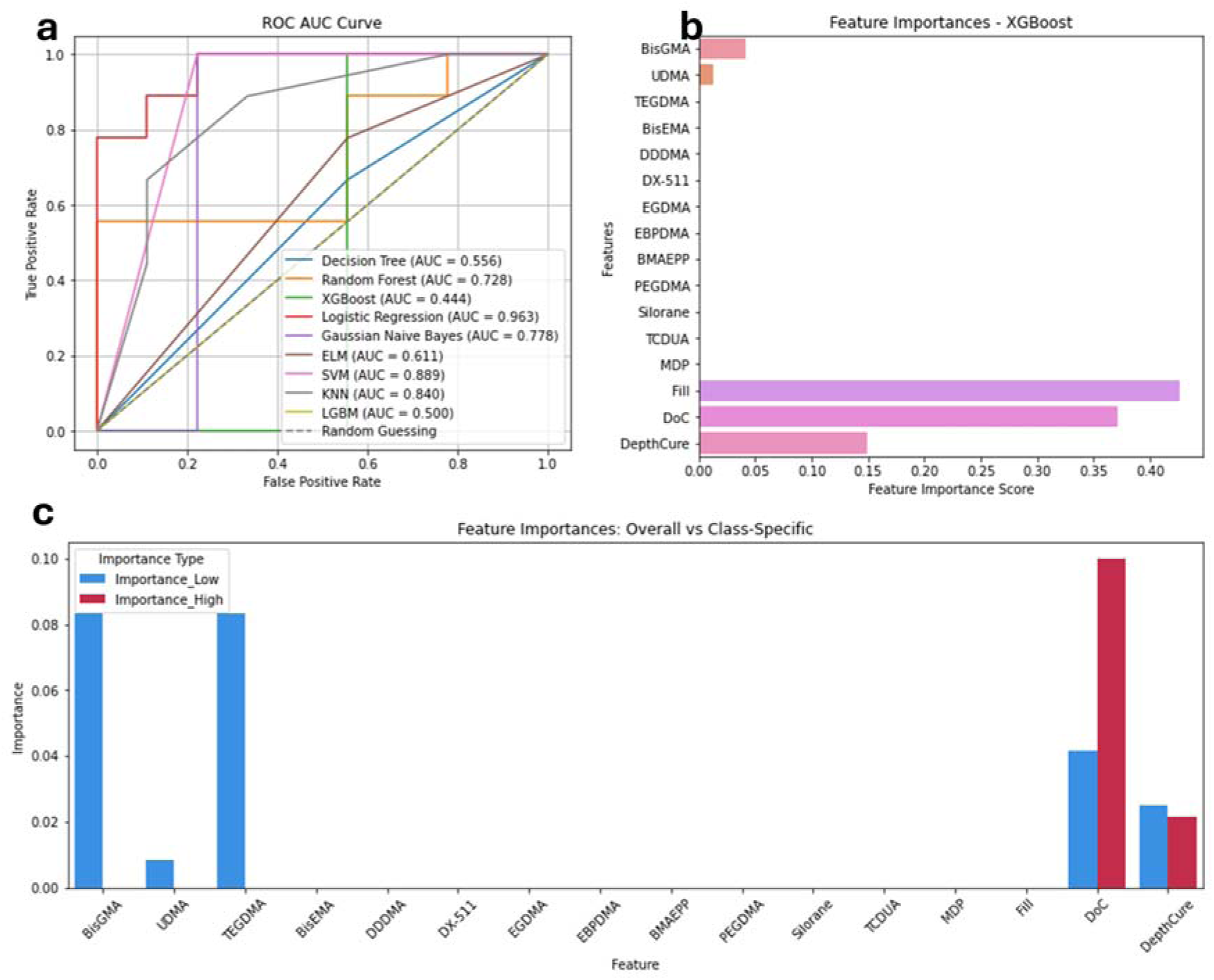
Shrinkage Stress. a) ROC AUC Curve of the 9 algorithms, b) Feature importances for Shrinkage Stress. c) Feature importances per class.

Feature importance analysis measured using XGBoost (Figure 4b) shows that the six most important CAs for the prediction of ShrinkStr were, in order of highest to lowest importance: Fill (FIS = 0.43), DoC (FIS = 0.37), DepthCure (FIS = 0.15), BisGMA (FIS = 0.04), and UDMA (FIS = 0.01). The other CAs had negligible importance.

Figure 4c shows the importance of the CAs in predicting low or high ShrinkStr. For low ShrinkStr, BisGMA and TEGDMA were the most important features (PS = 0.08 for both CAs), followed by DoC (PS = 0.04), DepthCure (PS = 0.03) and UDMA (PS = 0.008). For high ShrinkStr, DoC was the most important feature (PS = 0.10), followed by DepthCure (PS = 0.02).

### 3.3. Regression

Table 9 shows the R^2^ scores for each of the analyzed POs. The Voting Regressor model demonstrated superior performance in predicting ShrinkV and FlexMod, achieving R^2^ scores of 0.83 and 0.91, respectively. In contrast, the Decision Tree Regression model was best for predicting FlexStr and ShrinkStr, with R^2^ scores of 0.82 and 0.93, respectively.

**Table 9.** R^2^ measure for regression.

| Model | FlexMod | FlexStr | ShrinkV | ShrinkStr |
| --- | --- | --- | --- | --- |
| SVR | 0.83 | 0.32 | 0.72 | 0.77 |
| Decision Tree Regression | 0.77 | **0.82** | 0.74 | **0.93** |
| HistGradientBoostingRegressor | 0.84 | 0.70 | 0.79 | -0.21 |
| Random Forest Regressor | 0.77 | 0.41 | 0.77 | 0.84 |
| Voting regressor | **0.91** | 0.64 | **0.83** | 0.91 |

Figure 5 confirms the R^2^ results showing how well the actual values match the predicted values. Most of the data points are clustered closely around the ideal (dotted) line, indicating that the predicted values are very close to the actual values for most instances for the four POs analyzed.

**Figure 5.**
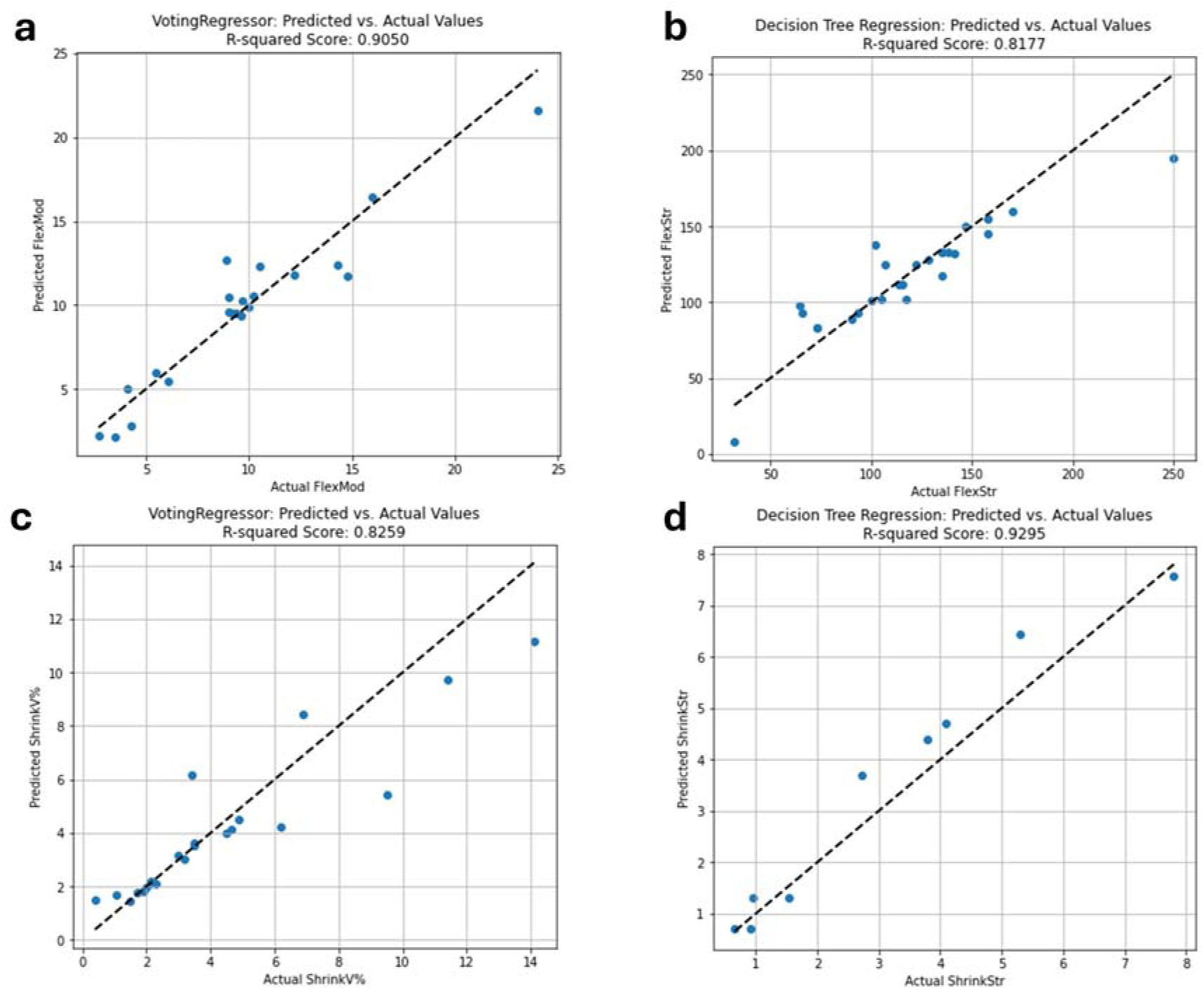
Model fit: Predicted vs Actual Values from a) Voting Regressor for FlexMod, b) Decision Tree for FlexStr, c) Voting Regressor for ShrinkV, and d) Decision Tree for ShrinkStr.

Table 10 shows the errors of the best regression models used to predict the different POs to assess the models’ performances. Overall, ShrinkStr has the lowest error metrics with MAE = 0.49 MPa, MSE = 0.36 MPa, RMSE = 0.6 MPa, MAE = 0.34 MPa, and ME = 1.15 MPa, indicating that the model predicts ShrinkStr values close to the actual measurements. However, for FlexStr, despite achieving a relatively high R^2^ score of 0.82, the error metrics had the highest discrepancies between predicted and actual values, underscoring the challenges in accurately predicting FlexStr using the current models. Model refinement or alternative approaches may be necessary to improve prediction accuracy. For algorithms with lower performance, detailed error scores are provided in Supplemental Tables 1-4.

**Table 10.** Error of the Best Regression Models.

| Error | FlexMod:<br>Voting regressor<br>(GPa) | ShrinkV:<br>Voting<br>regressor (%) | FlexStr:<br>Decision Tree<br>Regression<br>(MPa) | ShrinkStr<br>Decision Tree<br>Regression (MPa) |
| --- | --- | --- | --- | --- |
| Mean Absolute Error | 1.08 | 0.87 | 11.86 | 0.49 |
| Mean Squared Error | 2.17 | 2.01 | 318.22 | 0.36 |
| Root Mean Squared Error | 1.47 | 1.42 | 17.83 | 0.6 |
| Median Absolute Error | 0.55 | 0.28 | 7.19 | 0.34 |
| Max Error | 3.77 | 4.07 | 55 | 1.15 |

Feature importance analysis using the regression models for each of the four predicted POs showed that for FlexMod, TEGDMA (FIS = 0.68), DoC (FIS = 0.15), and Fill (FIS = 0.13) were the most important (Figure 6a). For FlexStr, the most important CAs were UDMA (FIS = 0.33), TEGDMA (FIS = 0.23), BisGMA (FIS = 0.17), DepthCure (FIS = 0.11), DoC (FIS = 0.09) and Fill (FIS = 0.04) (Figure 6b). For ShrinkV, the most important features were TEGDMA (FIS = 0.52), DoC (FIS = 0.24), DepthCure (FIS = 0.18), BisGMA (FIS = 0.04), Fill (FIS = 0.01) and UDMA (FIS = 0.01) (Figure 6c). For ShrinkStr, the most important features were DepthCure (FIS = 0.57), DoC (FIS = 0.30), Fill (FIS = 0.12) and BisGMA (FIS = 0.007) (Figure 6d).

**Figure 6.**
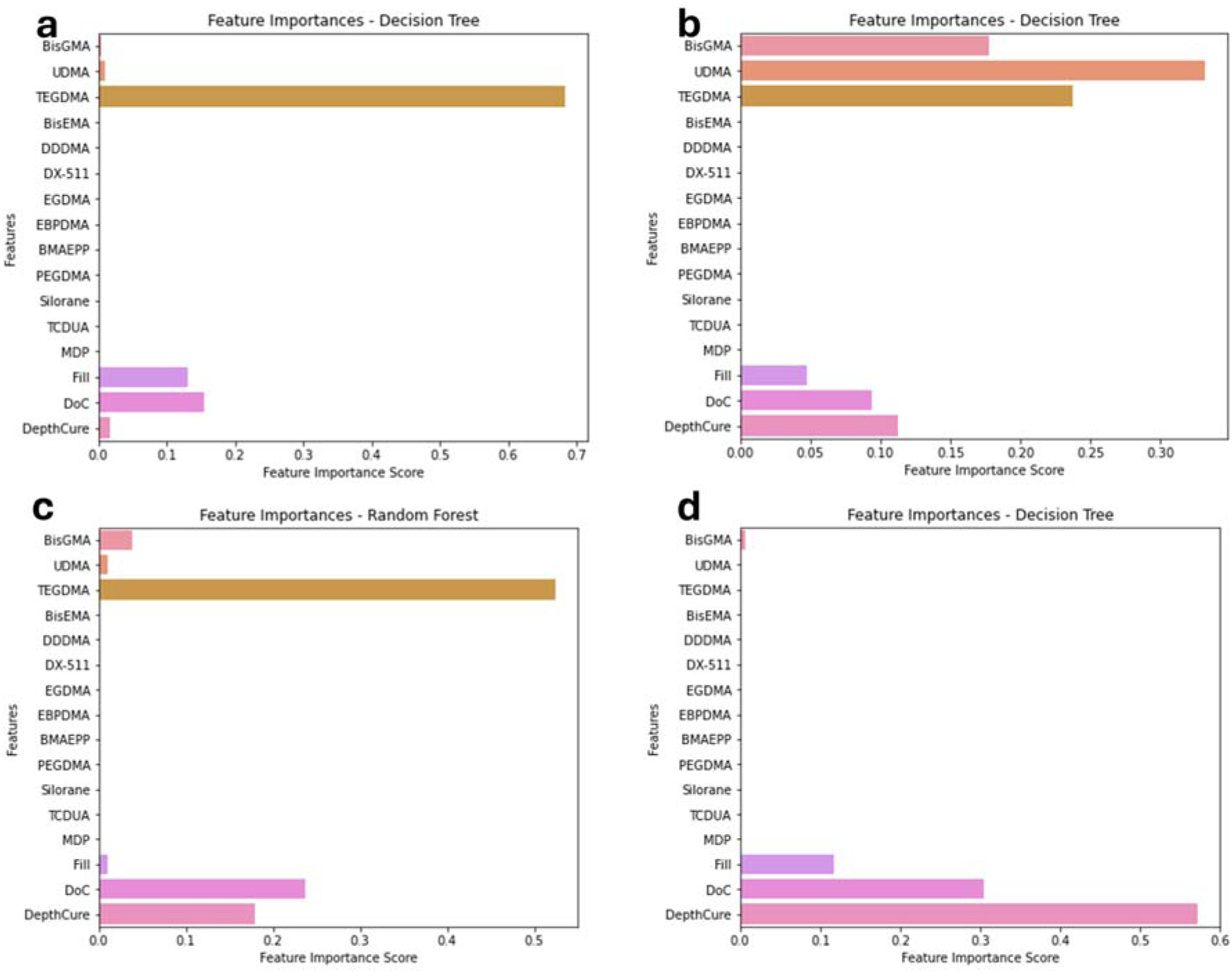
Feature importances for a) FlexMod, b) FlexStr, c) ShrinkV, and d) ShrinkStr.

## 4. DISCUSSION

The specific objectives of this work were to build a larger dataset of composite attributes (CAs) and composite performance outcomes (POs) from an extensive literature search of dental composites and use that to determine the efficacy of different machine learning (ML) models in predicting POs so we can optimize composite design and develop durable composites. Despite the use of 200+ publications and gathering data for 321 composites, there were many empty cells due to the different data that was presented and the fact that much of the dental composites literature is focused on novel monomers and filler systems that are not widely used. Thus, only data on commercial composites were used, and after processing, a final dataset of 233 composite samples consisting of 17 CAs and 7 POs were analyzed with different ML models.

As mentioned above, the only other published article on using AI to predict POs at the time of the writing of this manuscript only contained 12 samples, and most of that data was derived from company brochures [20]. While the results seemed to be very good, there is a danger of overfitting due to the small sample size. This was also evident in the present study for the results on compressive strength, fracture toughness and fracture work, which had perfect AI performance scores, but probably due to small samples sizes and overfitting. Thus, these Pos were deemed unreliable and excluded from further analyses. Furthermore, it was found that the reported PO data on the same commercial composite differed markedly from publication to publication, probably due to the different methods and instruments used to determine the POs. Thus, the first part of this work was a first attempt at gathering this data. Nonetheless, as can be seen from the POs that were excluded from all the analyses and the values that needed to be imputed, there is a need for more complete data on dental composites. Unlike in other fields and applications of AI, where there are tens of thousands of datasets, that is not the case here.

In addition, the current dataset needs to be expanded to include data from experimental composites. The difficulty with this is that those composites tend to use unique monomers or fillers, leading to small sample sizes and making their contribution to POs more difficult to determine. Regardless, this is a necessary step if novel composites are to be developed. Thus, a future goal of the authors is to create an opensource dataset, where researchers can contribute their research data in exchange for the use of the dataset for their research. Another potential way to mitigate the small sample set for unique monomers and fillers, and a future goal, is to use chemical formulas as CAs instead of simply monomer name and/or concentration.

For classified data (contribution to above average or below average POs), KNN performed the best in predicting low or high flexural modulus (FlexMod, Table 3 and Fig. 1a), Decision Tree performed the best for flexural strength (FlexStr, Table 4) and shrinkage volume (ShrinkV, Table 7), and Logistic Regression and SVM performed the best for shrinkage stress (ShrinkStr, Table 8). KNN’s superior performance for classifying FlexMod (Table 3 and Fig.1a), can be attributed to two factors: 1) KNN is good at identifying local patterns within the data. This is done by evaluating how similar the CA values are for two or more samples that belong to the same class, or the proximity of CA data points in the same PO class. Thus, it is called Nearest Neighbors. Despite this, KNN may not outperform other models for all the POs, as the effectiveness of each model can depend on the specific characteristics and relationships between each PO and the CAs.

2) Unlike linear models, such as Logistic Regression and Gaussian Naïve Bayes, KNN is a non-parametric model. This means it does not assume a specific form of relationship between features (CAs) and the target variable (PO), and that allows it to handle complex, non-linear relationships without needing prior knowledge about the data distribution [34]. This flexibility becomes evident when comparing KNN’s performance metrics with those of linear models (Table 3). For instance, KNN achieved a higher accuracy (0.90), precision (0.92), recall (0.90) and F1 score (0.90) compared to Logistic Regression (accuracy: 0.83, precision: 0.85, recall: 0.83, F1 score: 0.82) and Gaussian Naïve Bayes (accuracy: 0.73, precision: 0.77, recall: 0.73, F1 score: 0.73). Thus, KNN can better identify local patterns when handling complex, non-linear relationships in the data.

On the other hand, Decision Tree performed best in predicting low or high FlexStr and ShrinkV. Decision Tree is based on classification trees, a tool that uses a tree-like model of decisions and their possible consequences. It splits the data into subsets based on the values of input features, creating branches that lead to different outcomes or class labels [35]. However, despite achieving the highest accuracy, precision, recall and F1 scores, the Decision Trees AUC scores were not the highest (Figures 2a and 3a), probably due to the its tendency to overfit the training data, especially when the model becomes too complex. The classification tree for FlexStr had 31 nodes and a depth of 6, and the tree for ShrinkV had 21 nodes and depth of 9 (Figures S2 and S3) [36], which may be too complex for the data size. This complexity can lead the model to overfit the data, i.e. memorize the training data and perform very well on the training data but struggle to generalize to new data because it has memorized instead of identifying patterns. This can affect the model’s ability to distinguish between low and high FlexStr and ShrinkV across different AUC decision thresholds, affecting the AUC score. This inconsistency in ranking means that the model struggles to maintain a high true positive rate (real high FlexStr values correctly predicted) across various AUC decision thresholds, leading to a lower AUC score.

Alternatively, ensemble methods, like Random Forest and XGBoost, aggregate predictions from multiple decision trees [37], allowing them to identify more relationships within the data and produce predictions that are more robust across a range of AUC decision thresholds. Specifically, Random Forest builds multiple trees using different subsets of the data and features, which reduces variance and improves generalization [30]. XGBoost enhances the performance by iteratively refining predictions through a series of models that focus on correcting previous errors, using gradient descent for optimization and incorporating regularization to prevent overfitting [38]. In contrast, the Gaussian Naïve Bayes model, even though it is simpler, assumes independence between features which works well when the data (CAs and PO) is not too complex [39].

Despite this, the Decision Tree remains a good choice. Although it has a lower AUC score, it achieved higher performance metrics (accuracy, precision, recall, and F1 score), and it is highly interpretable. This interpretability comes from the tree’s structure, where each decision node represents a feature split based on a certain threshold, and the branches represent different outcomes or decisions (Figures S2 and S3). This structure allows users to trace the path from features to final class labels, making it easy to understand how the predictions are made. For example, for FlexStr we can see that BisGMA was the most influential feature for classification, and how it interacts with the other CAs to lead to the model’s final prediction (Figure S2). For ShrinkV we can see that TEGDMA was the most influential feature for classification (Figure S3).

For ShrinkStr, the Logistic Regression, SVM and XGBoost models had high performance metrics (Table 8). However, only Logistic Regression and SVM models had high AUC scores (Figure 5a). Logistic Regression stands out in its probabilistic interpretation and simplicity, making it well-suited for tasks where understanding the impact of individual features is crucial, since it calculates the probability of an individual event happening [40]. SVM’s strength lies in its ability to handle complex, non-linear relationships through kernels, by mapping data into higher-dimensional spaces for better separation of classes [41]. XGBoost’s lower AUC score suggests challenges with generalization or overfitting, despite its robust overall performance.

Finally, in the regression analysis, the Voting Regressor, an ensemble approach, benefited from combining multiple models’ predictions, leading to robust performance across FlexMod and ShrinkV, which may involve complex interactions among features (Table 9, Figure 5). On the other hand, the Decision Tree Regression demonstrated its capacity to capture complex relationships when predicting FlexStr and ShrinkStr (Table 9, Figure 5), because Decision Trees are good at modeling non-linear relationships and interactions between CAs.

The variability in model performance across different POs can be attributed to the specific characteristics of each PO and how they relate to the CAs. For instance, POs with more straightforward or linear relationships with the CAs may benefit from the probabilistic and direct nature of models like the Voting Regressor. In contrast, POs that involve more complex interactions might fare better with models that can handle non-linear data, such as the Decision Tree Regression. This comprehensive analysis (performance metrics and error rates) highlights the efficacy of ensemble methods like the Voting Regressor for linear and generalizable predictions (Tables 9 and 10), and the predictive power of Decision Tree Regression for capturing complex interactions across CAs within dental composite materials (Tables 9 and 10).

Our study’s results for FlexStr demonstrate greater robustness and reliability compared to the findings of Li et al. [20], primarily due to the significantly larger sample size used in our analysis (128 samples versus 12 samples in their study) improving the ability to detect patterns and relationships within the data. A larger sample size generally provides a more accurate and generalizable representation of the data, enhancing the validity of the results [42]. Despite this, for both studies, the highest R^2^ scores are achieved by Decision Tree based algorithm. In contrast, the smaller sample size in Li et al.’s study may limit the generalizability of their findings and increase the risk of overfitting.

In short, the evaluation of the predictive capabilities of various ML models for dental composite properties underscores the importance of dataset size and model selection in achieving accurate and reliable predictions for PO. One model does not fit all the POs best because the relationship between CAs and POs may or may not be linear. Thus, as the dataset grows, and the results become more robust, different models may need to be used to best predict composite POs.

Table 11 shows a summary of results of the feature importance analyses using classified data and regression analysis. Feature importance analysis demonstrated some consistency between the regression and classification models. First, both analyses identified TEGDMA as the most important feature for FlexMod (Table 11, Figure 1b, 6a) and for ShrinkV (Table 11, Figure 3b, 6c). These results are plausible. Due to TEGDMA’s small molecular weight, increase in TEGDMA concentration would increase crosslink density and increase composite modulus. However, it is interesting that filler loading did not have more of an effect, since inorganic filler has higher modulus than resins. TEGDMA’s importance in polymerization shrinkage volume is also logical, as its low molecular weight is the main cause of increased polymerization shrinkage. Interestingly, it is not listed as a major contributor to shrinkage stress, since increased shrinkage generally leads to increased shrinkage stress. Nonetheless, composites with higher depth of cure and higher filler loading tend to be in low-shrinkage bulk cure composites, so that may have overshadowed the effect of TEGDMA.

**Table 11:** Comparison of Important CAs in Feature Importance Analysis.

| FlexMod |  | FlexStr |  | ShrinkV |  | ShrinkStr |  |
| --- | --- | --- | --- | --- | --- | --- | --- |
| Classified Data | Regression Analysis | Classified Data | Regression Analysis | Classified Data | Regression Analysis | Classified Data | Regression Analysis |
| TEGDMA | TEGDMA | BisGMA | UDMA | TEGDMA | TEGDMA | BisGMA | DepthCure |
| BisGMA | DoC | DepthCure | TEGDMA | BisGMA | DoC | DepthCure | DoC |
| Fill | Fill | DoC | BisGMA | Fill | Fill | DoC | Fill |
| DepthCure | DepthCure | Fill | DepthCure | DepthCure | DepthCure | Fill | BisGMA |
| DoC | UDMA | UDMA | DoC | DoC | UDMA | UDMA |  |
| UDMA |  | TEGDMA | Fill | UDMA |  | TEGDMA |  |

For FlexMod, both analyses also identified Fill, DepthCure, DoC and UDMA as important features, although not in the same order of importance (Table 11, Figure 1b, 6a). For FlexStr, BisGMA, DepthCure, DoC, Fill, UDMA, and TEGDMA were recognized as important features in both analyses, but not in the same order of importance (Table 11, Figure 2b, 6b). BisGMA (most important feature for classification) and UDMA (for regression) are usually the largest and strongest components of the monomer system, so it is not surprising that it is recognized as a main contributor to FlexStr. Both analyses of ShrinkV also identified Fill, DepthCure, DoC and UDMA as important features, in different order of importance (Table 11, Figure 3b, 6c). And both analyses of ShrinkStr identified DepthCure, DoC, Fill and BisGMA as important features in different order of importance (Table 11, Figure 4b, 6d). Again, DepthCure may be identified as important due to the low shrinkage and shrinkage stress of bulk fill composites overshadowing the effects of other CAs, and it is known that high filler loading is important in reducing shrinkage and shrinkage stress. It is interesting that UDMA and BisGMA were not identified as more important in the regression analysis, even though BisGMA was identified as one of the most important CAs.

However, there were also discrepancies between the classification and regression analyses. BisGMA was identified as the second most important feature for FlexMod classification but was not identified as an important feature in regression. For ShrinkV, DepthCure was identified as the third most important feature in regression, while for classification it was not recognized as an important feature. Finally, for ShrinkStr, UDMA was identified as the least important feature for ShrinkStr, but still considered important, for classification analysis, but it was not recognized as important for the regression analysis. These discrepancies are due to the two models having different goals. While regression is trying to estimate a PO value from the CA values, classification is just indicating if the PO will be low or high. Thus, some difference is expected.

Finally, while filler loading should have also been a major contributor, it is possible that most of the samples were filled and hence, its variability is limited, and its effect underestimated. Furthermore, sometimes highly filled composites can be more brittle and lead to lower flexural strength.

Our results indicate a need to create a massive database of commercial and experimental CAs and POs that our AI models can use to better predict POs for new dental composites and predict optimal CAs needed to produce specific POs. With more data, optimized AI models would allow dental materials researchers and manufacturers to input the CAs of their experimental composites and get a prediction of various POs before they fabricate and test their first composite. Our present work consists in the development of such a tool. This will significantly reduce development time because it would narrow the starting groups for testing.

## 5. CONCLUSIONS

An extensive database of 321 dental composites with 28 composite attributes (CA) and 17 performance outcomes (PO) was built using 200+ publications. However, many CA and PO values were missing due to a lack of data in the literature. This data was curated down to 233 composites with 17 CAs and 7 POs. A larger database is needed to train more effective AI models. This dataset was preprocessed and used to train and test nine machine learning (ML) models to predict POs using the CAs. For classified data, the KNN model performed the best in predicting flexural modulus (FlexMod), the Decision Tree model performed the best for flexural strength (FlexStr) and volumetric shrinkage (ShrinkV), and the Logistic Regression and SVM models performed the best for classifying shrinkage stress (ShrinkStr). Receiver Operating Characteristic Area Under the Curve (ROC AUC) analysis concurred with the KNN, Logistic Regression and SVM results but found the Random Forest model to be more effective for FlexStr and ShrinkV, possibly due to overfitting by the Decision Tree model. For continuous data, regression analysis showed that the Voting Regressor was best for FlexMod and ShrinkV, and the Decision Tree Regression was best for predicting FlexStr and ShrinkStr. One model does not work best for all POs and need to be selected based on full analysis. Feature importance analysis on classified data showed some consistency to both the classified and regression analyses. For FlexMod and ShrinkV, TEGDMA contributed the most, for FlexStr, BisGMA and UDMA contributed the most, and for ShrinkStr, depth of cure and degree of monomer-to-polymer conversion were the most important CAs. These CAs need to be controlled carefully to produce composites with the desired POs. AI can be used to predict composite POs based on CAs and be used to design dental composites, but the right model needs to be chosen and a more extensive database of CAs and POs is needed.

## Data Availability

All data produced in the present study are available upon reasonable request to the authors

## 6. ACKNOWLEDGEMENTS

This work was supported by National Institutes of Health grants NIDCR DE031477 (Y.S.K.), and NS128574 (Y.S.K.), Rising STAR Award (Y.S.K.) from the University of Texas System. NIH Artificial Intelligence/Machine Learning Consortium 1OT2OD032581-1-24 (M.F.) and HUB SPECIFIC PILOT 1OT2OD032581-1-31 (M.F)

## Supplementary file

**Table S1.** Error scores for Shrinkage Volume.

| Error | SVR | Decision Tree Regression | Hist Gradient Boosting Regressor | Random Forest Regressor | Voting regressor |
| --- | --- | --- | --- | --- | --- |
| Mean Absolute Error | 1.24 | 0.85 | 0.89 | 1.07 | 0.87 |
| Mean Squared Error | 3.23 | 3.06 | 2.4 | 2.65 | 2.01 |
| Root Mean Squared Error | 1.79 | 1.75 | 1.55 | 1.63 | 1.42 |
| Median Absolute Error | 0.73 | 0.13 | 0.46 | 0.62 | 0.28 |
| Max Error | 5.66 | 5.29 | 5.4 | 5.18 | 4.07 |

**Table S2.**
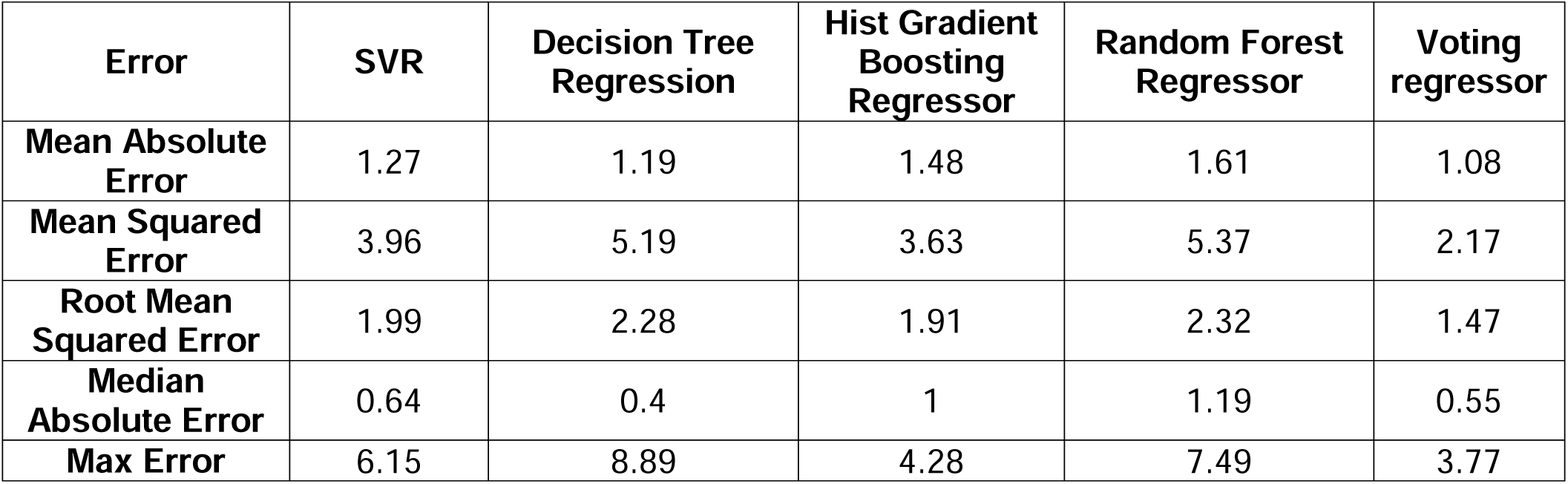
Error scores for Flexural Modulus.

**Table S3.** Error scores for Flexural Strength.

| Error | SVR | Decision Tree Regression | Hist Gradient Boosting Regressor | Random Forest Regressor | Voting regressor |
| --- | --- | --- | --- | --- | --- |
| Mean Absolute Error | 23.14 | 11.86 | 15.32 | 22.87 | 17.02 |
| Mean Squared Error | 1180.08 | 318.22 | 521.12 | 1028.27 | 626.41 |
| Root Mean Squared Error | 34.35 | 17.83 | 22.82 | 32.07 | 25.03 |
| Median Absolute Error | 11.39 | 7.19 | 6.05 | 16.59 | 10.22 |
| Max Error | 98.64 | 55 | 0.7 | 119.46 | 93.18 |

**Table S4.**
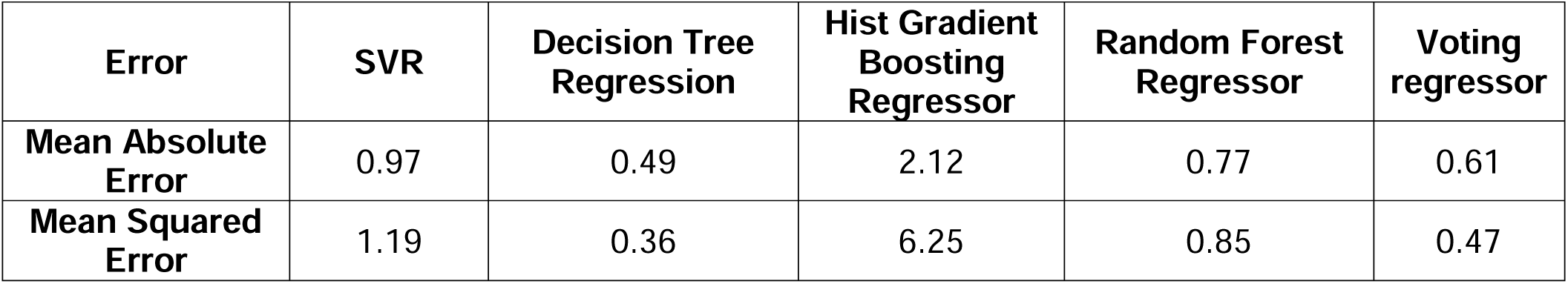
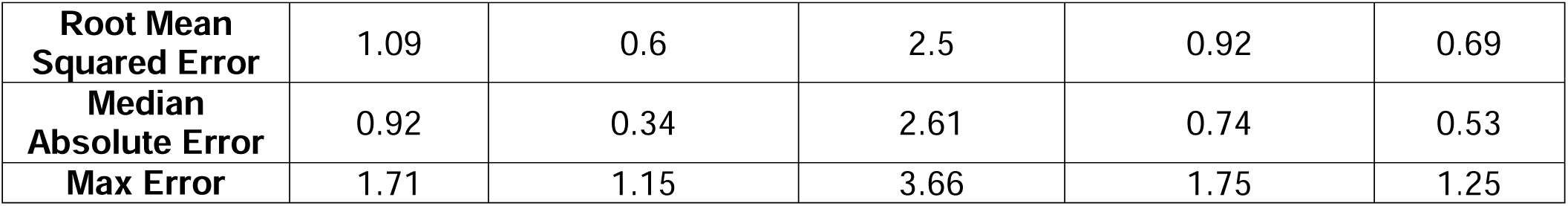
Error scores for Shrinkage Stress.

| Error | SVR | Decision Tree Regression | Hist Gradient Boosting Regressor | Random Forest Regressor | Voting regressor |
| --- | --- | --- | --- | --- | --- |
| Mean Absolute Error | 0.97 | 0.49 | 2.12 | 0.77 | 0.61 |
| Mean Squared Error | 1.19 | 0.36 | 6.25 | 0.85 | 0.47 |
| **Root Mean Squared Error** | 1.09 | 0.6 | 2.5 | 0.92 | 0.69 |
| **Median Absolute Error** | 0.92 | 0.34 | 2.61 | 0.74 | 0.53 |
| **Max Error** | 1.71 | 1.15 | 3.66 | 1.75 | 1.25 |

**Figure S1.**
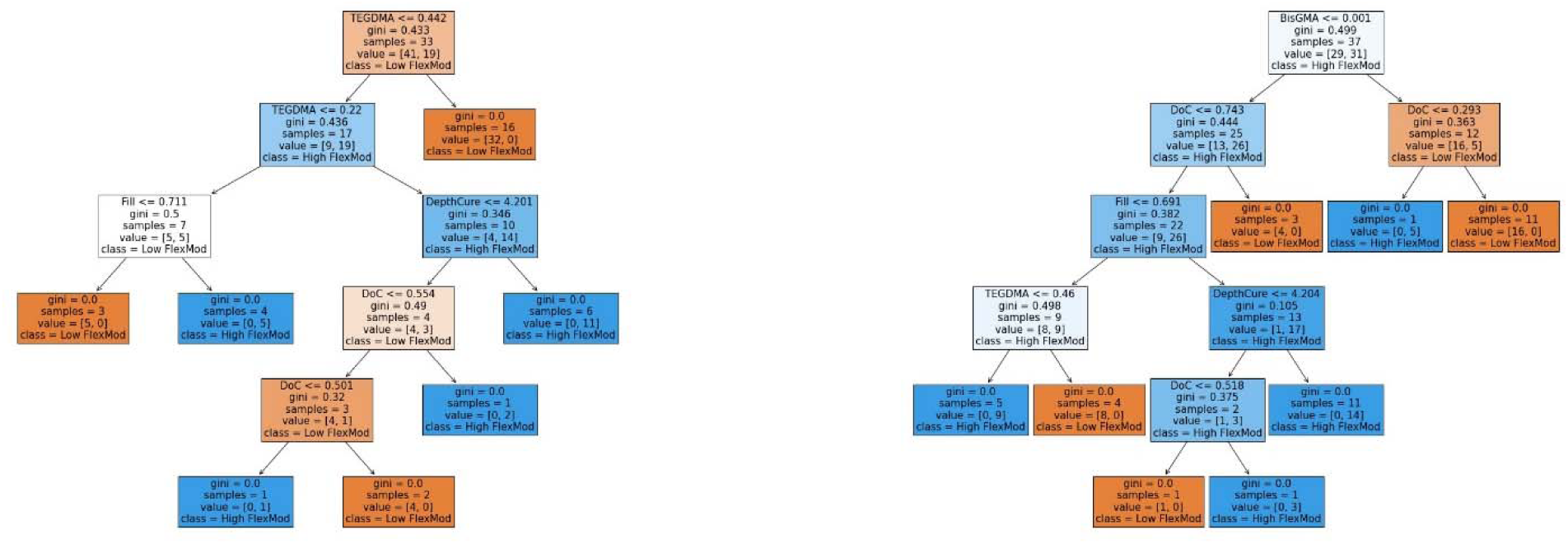
Random Forest of two the one hundred estimators, where gini is the impurity score.

**Figure S2.**
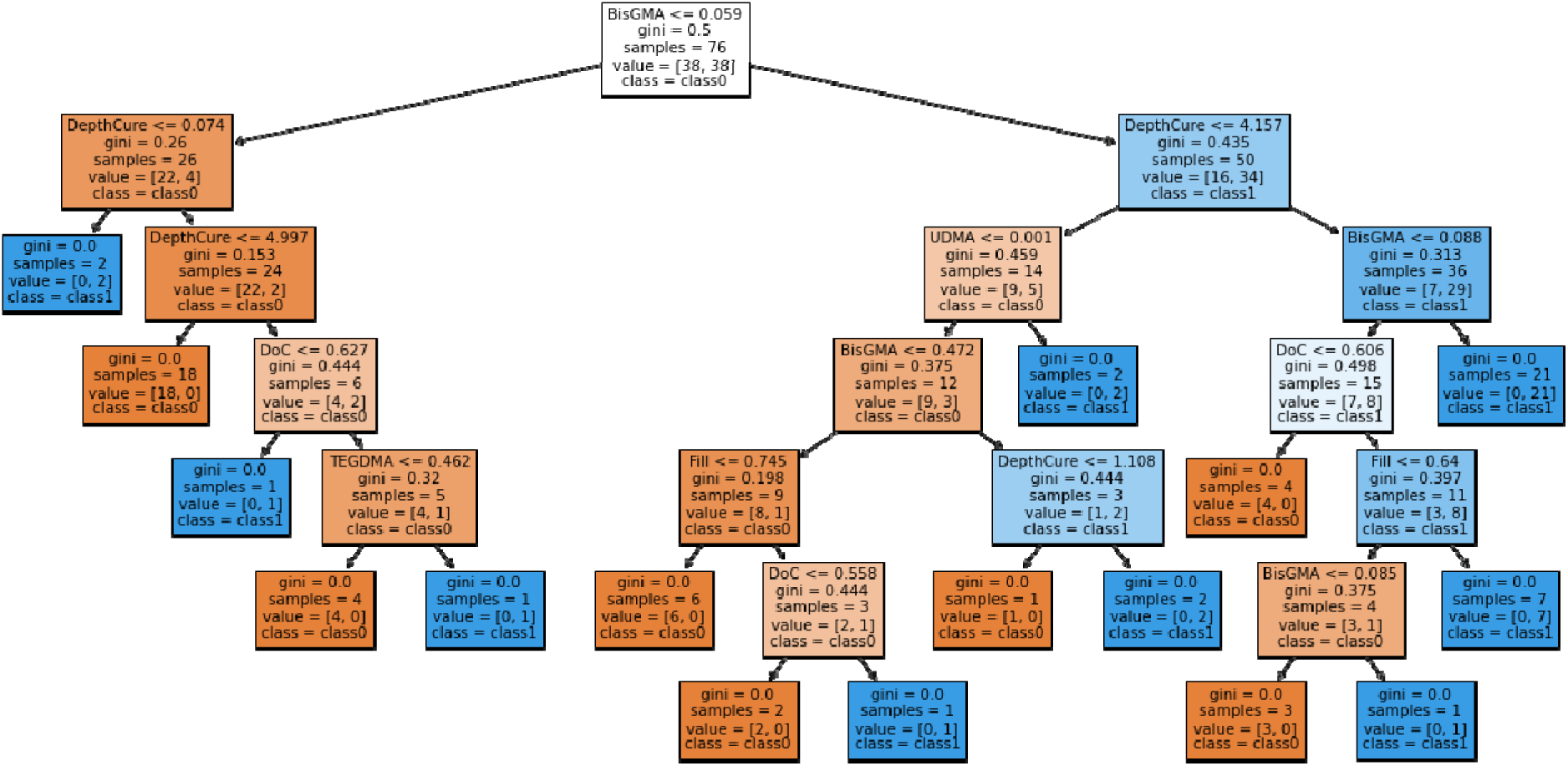
FlexStr Decision Tree. 31 nodes and Depth of 6.

**Figure S2.**
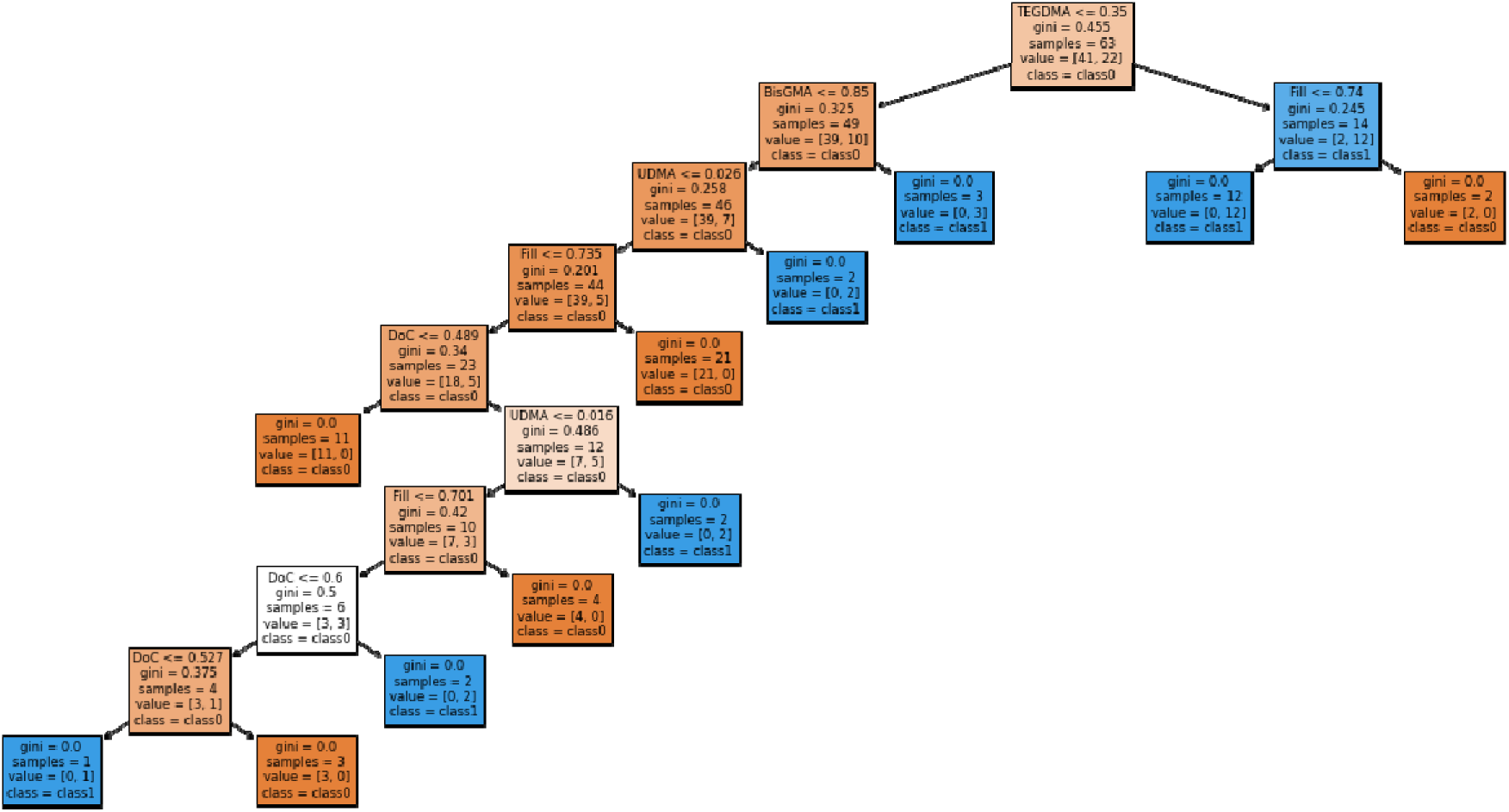
ShrinkV Decision Tree. 21 nodes and Depth of 9.

